# CALGARY NORMATIVE STUDY: STUDY DESIGN OF A PROSPECTIVE LONGITUDINAL STUDY TO CHARACTERIZE POTENTIAL QUANTITATIVE MR BIOMARKERS OVER THE ADULT LIFESPAN

**DOI:** 10.1101/2020.02.28.20028894

**Authors:** Cheryl R McCreary, Marina Salluzzi, Linda B Andersen, David Gobbi, M Louis Lauzon, Feryal Saad, Eric E Smith, Richard Frayne

## Abstract

**Introduction:** A number of magnetic resonance (MR) imaging methods have been proposed to be useful, quantitative biomarkers of neurodegeneration in aging. The Calgary Normative Study (CNS) is an ongoing single-centre, prospective, longitudinal study that seeks to develop, test and assess quantitative MR methods as potential biomarkers. The CNS has three objectives: first and foremost, to evaluate and characterize the dependence of the selected quantitative neuroimaging biomarkers on age over the adult lifespan; secondly, to evaluate the precision, variability and repeatability of quantitative neuroimaging biomarkers as part of biomarker validation providing proof of-concept and proof-of-principle; and thirdly, provide a shared repository of normative data for comparison to various disease cohorts.

**Methods and Analysis:** Quantitative MR mapping of the brain including longitudinal relaxation time (T1), transverse relaxation time (T2), T2*, magnetic susceptibility (QSM), diffusion and perfusion measurements, as well as morphological assessments are performed. The Montreal Cognitive Assessment (MoCA) and a brief, self-report medical history will be collected. Mixed regression models will be used to characterize changes in quantitative MR biomarker measures over the adult lifespan. In this report on study design, we report interim prevalence and demographic information of recruitment from 28 May 2013 to 31 December 2018.

**Ethics and Dissemination:** Participants provide signed informed consent. Changes in quantitative MR biomarkers measured over the adult lifespan as well as estimates of measurement variance and repeatability will be disseminated through peer-reviewed scientific publication.

**STRENGTHS AND LIMITATIONS:**

- Both cross-sectional and longitudinal quantitative MR data is being acquired in a large sample normal aging population to characterize changes in these potential neuroimaging biomarkers over the adult lifespan.
- Clinical and multiple quantitative imaging data are being collected and shared with the research community.
- Measures of repeatability and a process to update the imaging protocol are included in the study design.
- Associated self-reported medical history and cognitive assessments are limited

## INTRODUCTION

Quantitative imaging methods can be defined as “extraction and use of numerical or statistical features from medical images”.^1^ There are numerous examples where quantitative measures have improved detection of changes or monitoring of the brain.^2-4^ Specifically, quantitative neuroimaging using MR is thought to be suitable for identifying potential biomarkers of cognitive impairment risk because it 1) may be more sensitive to early detection of pathological changes within otherwise normal appearing tissue,^5-9^ 2) may be more directly related to underlying physiological processes of interest, 3) may have reduced variability across time and/or equipment platforms when compared to traditional, more qualitative, neuroimaging methods, 4) can provide less subjective interpretation, and 5) allows for a low risk, repeatable measurement.^1 10^

Numerous quantitative MR imaging biomarkers have been proposed to assess risk, progression and treatment of age-related neurodegenerative diseases and disorders including cerebral small vessel disease,^10^ Alzheimer’s disease,^3^ Parkinson’s disease,^11^ multiple sclerosis,^12 13^ and Huntington’s disease.^14^ These quantitative markers typically include measures of brain atrophy, white matter integrity, iron accumulation, and cerebral blood flow. Often, these measures can be determined using standard, vendor-provided MR sequences and freely available image processing packages. Less commonly, longitudinal (T1) and transverse (T2, T2*) relaxometry, vascular permeability and quantitative susceptibility mapping using more customized approaches have been used.

Development of quantitative imaging biomarkers requires appropriate validation and qualification as outlined by the Quantitative Imaging Biomarkers Alliance (QIBA) organized by the Radiological Society of North America.^15 16^ An advantage of quantitative imaging is that the measurements obtained should be, in principle, independent of the specifics of acquisition. While brain atrophy, white matter hyperintensity volume, white matter integrity as measured by diffusion imaging metrics, number of cerebral microbleeds, number and volume of infarcts (including ischemic, hemorrhagic, and lacunar), relative increase in tissue iron have all been associated with one or more neurodegenerative processes, detailed validation of these quantitative biomarkers is relatively lacking.^16^ Furthermore, quantitative methods like MR relaxometry, e.g., T1, T2, and T2* mapping, quantitative susceptibility mapping, and arterial spin labelling measurement of cerebral perfusion also require comprehensive evaluation or validation as potential quantitative neuroimaging biomarkers of neurodegenerative disease or pathology.

The Calgary Normative Study (CNS) is a single centre, prospective, longitudinal study with three objectives: 1) to evaluate and characterize the dependence of the selected quantitative neuroimaging biomarkers on age over the adult lifespan; 2) to evaluate the precision, variability and repeatability of quantitative neuroimaging biomarkers as part of biomarker validation providing both proof of-concept and proof-of-principle^16^; and 3) provide a shared repository of normative data for comparison to various disease cohorts. In developing this study, we developed a plan incorporate the ability to revise our data acquisition protocol so that we can refine and accommodate emerging quantitative techniques. We also aimed to complete all imaging procedures and other study evaluations within 2 hours. Our primary research interest is in cerebral small vessel disease and this guided our selection of potential quantitative neuroimaging biomarkers, though in both theory and practice they have proven to be of broad application in other neuroimaging studies. Here we describe the study design and methods used in the CNS and report on recruitment and data sharing. Reporting the specific short- and long-repeatability or changes in quantitative metrics over the adult lifespan are beyond the scope of this report and will be submitted as separate submissions for publication subsequently.

## MATERIALS AND METHODS

### Participant Eligibility and Characterization

This study was approved by the local research ethics board (REB). Study recruitment is ongoing; volunteers over 18 year of age are being recruited from the community, primarily through local poster advertisement and word-of-mouth. Interested individuals provided written informed consent and were screened for eligibility based on self-reported absence of significant neurological disease, psychiatric disorders, or contraindications for MR imaging at 3 T. Participants with uncertain contraindications (poorly characterized implant, *etc*.) were not imaged and excluded from the study. Participants provided a brief medical history, completed a Montreal Cognitive Assessment (MoCA)^17^, completed our institutional MR safety screening and underwent MR imaging.

Demographic and simplified medical information collected includes age, sex, ethnicity, handedness, years of education, smoking history, weight, hypertension or taking medication for the treatment of hypertension, dyslipidemia or taking medication for the treatment of high cholesterol, and presence of diabetes mellitus, and family history of stroke, dementia, Alzheimer’s disease, or cardiovascular disease. Participants were also asked if they are willing to return for a follow-up visit and if they are willing to be contacted in the future for possible participation in other research studies. All data were de-identified and labelled with CNS-specific identification numbers.

The MoCA is a cognitive screening test designed to assist the detection of mild cognitive impairment, which was administered and scored by trained research personnel. If participants were not fluent in English, then alternate language versions of the MoCA (Mandarin and Spanish) were offered and were administered with the aid of a translator. Participants that scored <26 on the MoCA were considered screen failures for normal cognition.^17^ However, these individuals still completed the study procedures, though we anticipate that their data may be excluded from some subsequent analyses. Participants were not informed of their MoCA scores because the MoCA alone is not sufficient to diagnose a clinical cognitive disorder such as MCI, and there will be false positive low scores. If concerns about a clinical cognitive disorder arose during the testing the participant was advised to consult their primary care practitioner.

We sought to enroll an approximately balanced distribution of men and women in each of six age categories: 18-29, 30-39, 40-49, 50-59, 60-69, 70+ years. We anticipate that recruitment of eligible normal older participants would be more difficult due to the increased prevalence of stroke and or other significant neurological disease with age. For this reason, we decided to group individuals 70 years of age and older into a single age category, anticipating that we would have difficulty recruiting a sufficient number of individuals over 80 years of age.

A nominal sample size of 20 men and 20 women was initially targeted for each age category, for a nominal CNS recruitment of 240. This sample size estimate was based on estimates of variance from previous work using voxel-based analysis of T2 changes in temporal lobe epilepsy.^2^ The study was initially powered (*α* = 0.05, 1-*β* = 0.8) to detect T2 changes of 3.4 ms or greater. Larger or smaller sample sizes may be required depending on the quantitative measure, region, and analysis method chosen. To accommodate differences in power requirements, protocol revisions, and secondary analyses (eg sex differences), recruitment will continue after the nominal sample size has been achieved. Establishing better variance estimates for each quantitative imaging method is one of the objectives of the Calgary Normative Study.

### MR Imaging Acquisition Protocol

MR imaging is being completed at a single centre on a 3 T MR scanner (MR750, General Electric Healthcare, Waukesha, WI) using the vendor-supplied, 12-channel head, neck and spine coil. Standard procedure at our centre is to perform daily quality control scans and to follow the vendor recommended service guidelines.

Prior to setting up the initial MR protocol, we informally surveyed five local research teams with respect to their MR imaging protocols and typical outcome measures. This survey guided some of our protocol decisions and ensured that aspects of the MR acquisition parameters were similar to other studies in our centre. This similarity in some protocol acquisitions makes it feasible to share and leverage the data appropriately for comparison with disease cohorts.

The MR imaging protocol includes conventional structural 3D T1-weighted (3D-T1), T2-weighted fluid attenuated inversion recovery (FLAIR), as well as diffusion, pseudo-continuous arterial spin labelling (pcASL), resting state functional MR imaging (rs-fMRI) based on blood oxygen level dependent (BOLD) contrast, quantitative susceptibility mapping (QSM), and both T1 (qT1) and T2 (qT2) relaxometry sequences. Susceptibility-weighted imaging (SWI) results were derived from the acquired QSM image data. The quantitative neuroimaging metrics of interest generated from each sequence are summarized in Table 1. Details of the acquisition protocol are summarized in Table 2. The field of view was 240 mm for all sequences except 3D-T1 and QSM, which have a field of view of 256 mm.

**Table 1:**
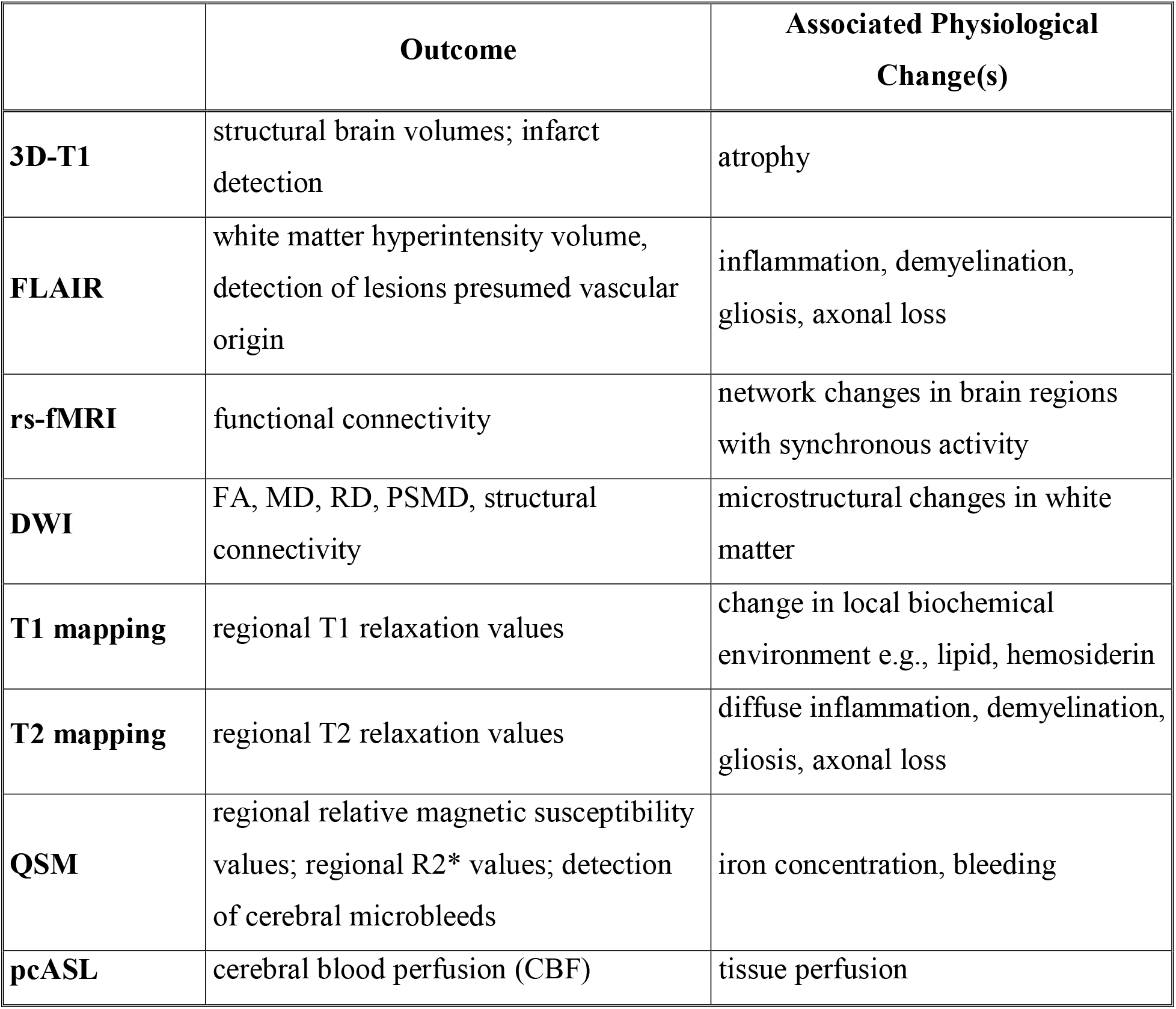
Acquired MR sequences, potential quantitative neuroimaging measurement outcomes, and associated physiological changes.

**Table 2A:** MR acquisition parameters for Phase 1 (*n*=54)

| **Sequence Type** | **Slice Orientation** | **TR (ms)** | **TE (ms)** | **Flip Angle (°)** | **Acquisition Matrix** | **Reconstructed Resolution (mm)** | **Other Parameters** | **Acquisition Duration (min:s)** |
| --- | --- | --- | --- | --- | --- | --- | --- | --- |
| **3D-T1** | coronal | ~7 | ~2.5 | 8 | 256 x 256 | 0.94 x 0.94 x 1.0 | TI = 650 ms | 5:03 |
| **FLAIR** | axial | 9000 | 148 | 90 | 256 x 256 | 0.94 x 0.94 x 3.0 | TI = 2250 ms | 4:50 |
| **rsfMRI** | axial | 2000 | 30 | 70 | 64 x 64 | 3.75 x 3.75 x 3.8 | 200 volumes | 5:10 |
| **DWI** | axial | 9000 | ~80 | 90 | 80 x 80 | 0.94 x 0.94 x 3.0 | $b=1000$ s/mm<sup>2</sup>; 31 directions | 6:01 |
| **qT1<sup>18</sup>** | axial | 15000 | ~24 | 40 | 160 x 160 | 1.0 x 1.0 x 4.0 | 8 echoes; B1 map included | 8:09 |
| **qT2** | coronal | 3000 | ~8-128 | 90 | 256 x 128 | 0.94 x 0.94 x 4.0 | 16 echoes; echo spacing = 8 ms | 9:48 |
| **QSM<sup>19</sup>** | axial | 30 | ~3-28 | 20 | 192 x 192 | 1.0 x 1.0 x 1.0 | 8 echoes; echo spacing = 3.4 ms | 4:13 |
| **pcASL** | axial | 4899 | ~11 | 111 | 512 x 10 | 1.88 x 1.88 x 5.0 | post label delay = 2025 ms; NEX = 1 | 6:55 |

**Table 2B:**
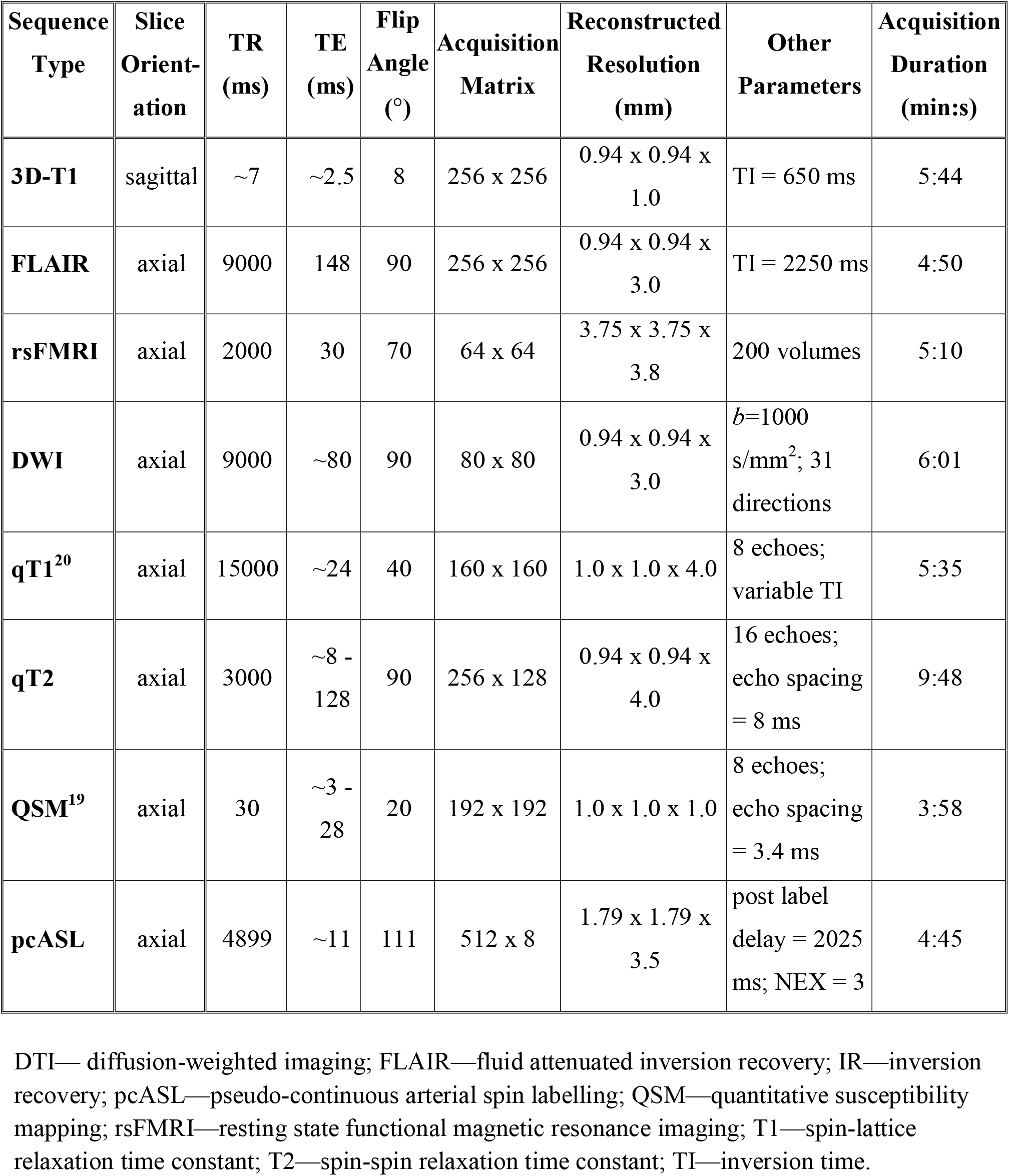
MR acquisition parameters for Phase 2 (*n*=271)

In designing this study, we recognized a need to evolve elements of the acquisition protocol, potentially in response to improved or new quantitative imaging techniques. The study was designed to incorporate distinct phases in order to accommodate protocol revisions. Revisions are proposed to and reviewed by the CNS protocol committee who evaluates the impact of the change, the number of participants completed in the current phase, and the number of overall protocol changes. The committee selects and recommends the appropriate timing of any proposed changes. This approach allows for a formal, orderly, well specified, evolution of the study data acquisition over time. To date, two study phases have been completed. The assigned CNS study identification number is used to help identify into which phase a subject is enrolled.

MR imaging was complete by trained staff (>15 years MR imaging research experience) or registered MR technicians. If any possible abnormalities in the images were noted, a neuroradiologist reviewed the images to determine if findings were of potential clinical significance, per the standard policy at our centre. Clinically significant findings as determined by the radiologist were reported to the participant by the radiologist following the procedure as outlined in our REB-approved ethics application.

### Image Assessment and Processing

Images were visually inspected for imaging artifacts like motion or spike noise, prior to any processing steps. The 3D-T1, FLAIR, diffusion-weighted and susceptibility-weighted images were reviewed more thoroughly to identify individuals with possible covert disease pathology associated with cerebral small vessel disease by an expert reader (FS) with >15 years of experience. Presence and number of lacunar stroke, cerebral microbleeds, recent subcortical infarcts, and scoring of white matter hyperintensities were recorded using the criteria for cerebral small vessel disease outlined in the STRIVE paper.^21^ Other incidental findings were noted and recorded. Standard and custom image processing pipelines will be used to generate and analyze the quantitative images and maps. These procedures are briefly summarized in Table 3.

**Table 3:**
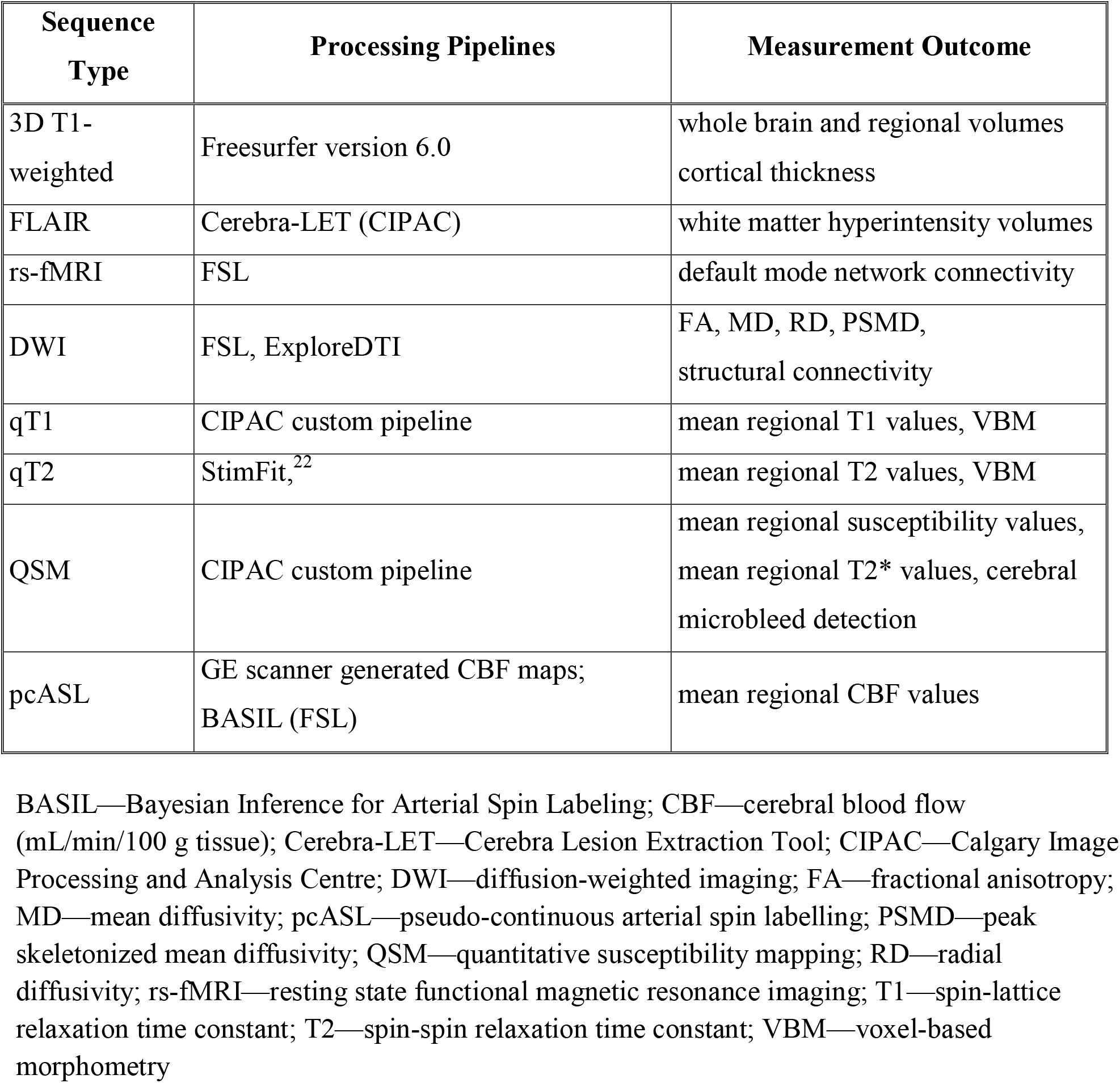
Image processing pipelines and measurement outcomes.

### Repeatability and Measurement Validation

A subset of four participants was asked at the time of the first scan to undergo repeated scanning for estimates of precision, variance and repeatability of the proposed quantitative neuroimaging biomarkers. Additionally, these data were used to determine if changes in the MR scanner software and hardware or in the MR acquisition parameters between protocol phases could be pooled or not depending on the sequence and processing pipelines used. Three additional time points were selected to include scanning before and after a MR system hardware and software upgrades. Not all sequences were acquired at all time points due to scanner and subject time availability. In these cases, a subset of data was acquired, which included 3D-T1, diffusion, T1 mapping, T2 mapping, ASL, QSM, but not FLAIR or rs-fMRI. Figure 1 illustrates the data acquisition timeline with scanner hardware or software changes.

**Figure 1:**
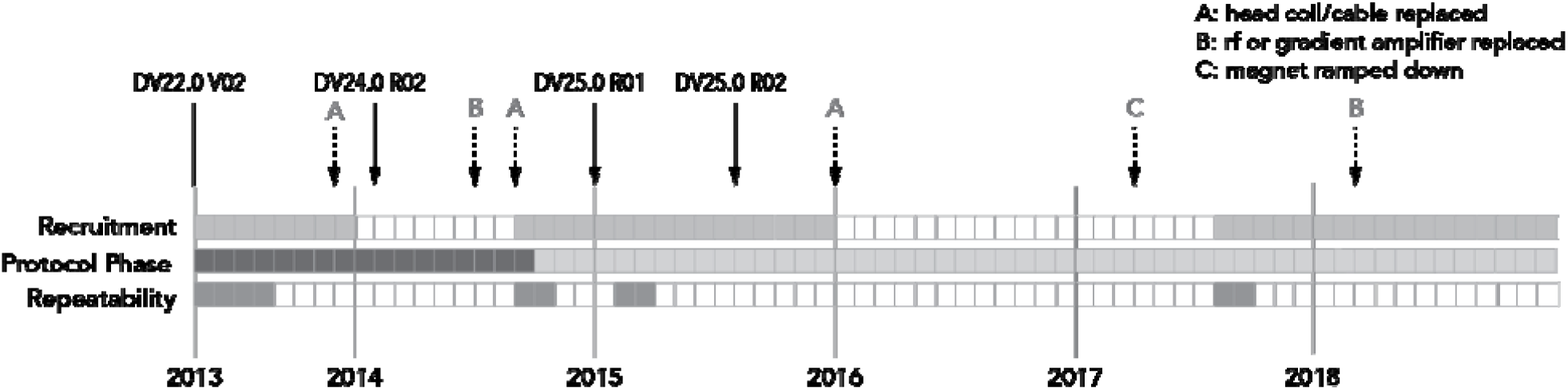
Timeline of MR protocols, recruitment phases, and scanner hardware and software upgrades. The filled squares, each representing one month, indicate when participants were actively recruited and scanned, when the MR acquisition protocol was revised and when repeatability measurements were completed. The solid arrows indicate version changes to the scanner software with the version number indicated above each arrow. The update between DV24.0 and DV25.0 included installation of some additional hardware. The dashed arrows indicate hardware repair or changes.

### Data Storage

De-identified demographic, medical, and cognitive assessment data were entered and maintained in a database (REDCap; Vanderbilt University, Nashville TN). MR image data were organized in an Osirix database where images can be reviewed clinically, or the database can be queried for specific criteria. Approximately 17 000 DICOM image files are acquired per imaging session requiring 1.3 GB of disk space for storage without compression, 510 MB with loss-less compression. The exact number of images for each individual can vary depending on head size and the number of slices required to cover the whole brain and whether sequences to be repeated due to motion, for example.

### Data Sharing, Ethics and Dissemination

MR image data and demographic information are available upon request for academic purposes from qualified researchers. Investigators interested in accessing these data are required to complete a data sharing agreement available on request by emailing. This brief, two-page form, requests a brief description of the study objectives and intended purpose, a list of MR sequences of interest, required medical information, and demographic information. The sharing agreement also requires that the study requesting data has approval from the appropriate REB. Once the agreement is approved by the CNS leadership and requesting principal investigators, the selected data are provided through secure electronic transfer. All data are de-identified and labelled with CNS-specific identification numbers. Groups requesting data sharing are asked to use the data for the specific intended purpose and to acknowledge the CNS and the CNS funding agencies.

Characterization of the changes in quantitative MR biomarker measures over the adult lifespan as well as estimates of measurement variance and repeatability will be disseminated through peer-reviewed scientific publication.

### Patient and Public Involvement

This research was done without patient or public involvement in the study design and they were not consulted to develop relevant outcomes or interpret the results. Patients were not invited to contribute to the writing or editing of this document for readability or accuracy.

## FINDINGS TO DATE

### Recruitment and Population Characteristics

Study enrolment is ongoing. This report describes the first 55 months of recruitment between 28 May 2013 and 31 December 2018. Over this period 325 participants have consented, provided study data, and completed at least one MR imaging session. In Phase 1 of the study, 54 subjects were recruited. Data from Phase 1 were reviewed and the ASL, T1, and T2 mapping sequences were modified prior to Phase 2 of the study. In the second phase, 271 subjects were recruited. Protocol changes are detailed in the following section “Protocol Revision”.

Additionally, as of 31 December 2018, 114 participants have returned for a second scan session after 41± 5 months (mean ± SD). Of the recruited participants, 24 (7.4%) were not interested in participating in other imaging studies and 33 (10.1%) were unsure or did not respond to the question prior to the first imaging session. Four individuals (1.2%) were no longer eligible for inclusion in the study after the baseline scan. Reasons for discontinuing participation in the study include existing diagnosis of multiple sclerosis (*n*=1) which was not disclosed until after the baseline MR scan, new diagnosis of mild cognitive impairment (*n*=1), new diagnosis of bipolar disorder (*n*=1) and multiple concussions (*n*=1).

The age and sex distributions of recruited participants are shown in Figure 2. Each age category has a nominal sample size of 40 individuals with nearly equal representation across the six age categories. More women than men (n=189 [58.1 %] versus n=136, respectively) have volunteered and this observation is consistent for each age category. Ninety percent of our participants were right-hand dominant. The distribution of represented ethnicities of the participants were 80.9% White, 16.6% Asian, 0.9% Hispanic, 0.3% First Nations, and 1.2% unknown or not reported.

**Figure 2A-D:**
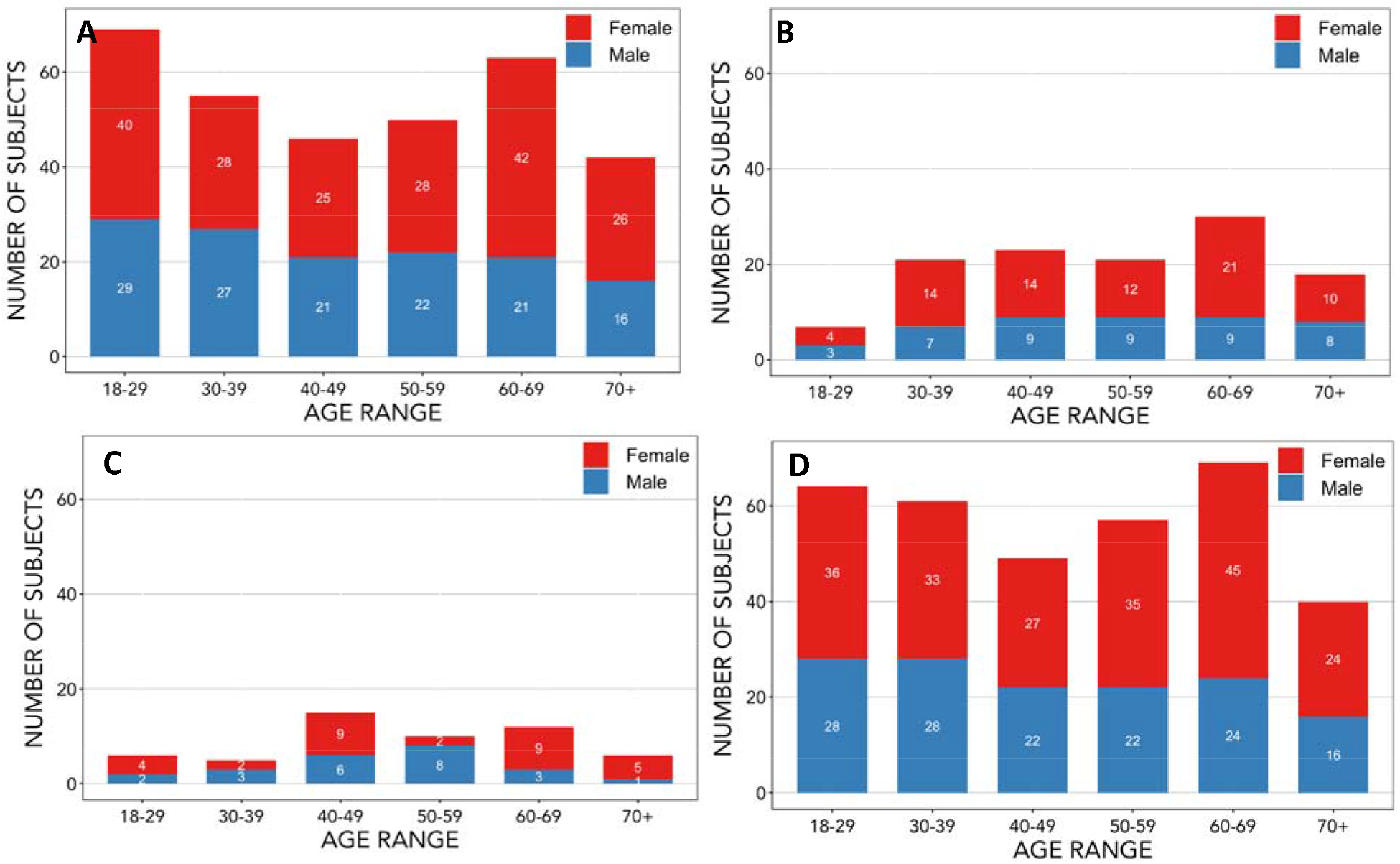
Distribution of age at study entry and sex of participants recruited up to December 2018. The age and sex distribution of participants has been provided for A) all baseline MR scans completed B) follow-up MR scans completed C) participants whose scans were completed with Phase 1 MR acquisition protocol and D) participants whose scans were completed with Phase 2 MR acquisition protocol. Phase 1 MR acquisition protocol was used only for baseline scans; Phase 2 MR acquisition protocol was used for some baseline and all follow-up scans. The exact number of participants for each sex in a given age category are indicated in the appropriate portion of the bar representing that category.

Neuroimaging indications of covert cerebral small vessel disease are summarized for each age category in Table 4. Other incidental findings that were noted either at the time of the scan by the scan operator or during review of the images by a qualified reader include: cysts (4 pineal, 3 subcutaneous dermoid, 1 arachnoid, and 2 other), 2 subcutaneous nodules, 2 megacisterna magna, 1 pituitary lesion, 1 venous angioma, 1 meningioma/lipoma, 1 asymmetrical cerebellar tonsils, and 1 empty sella tuncica.

**Table 4:** Study population prevalence of MR findings associated with cerebral small vessel disease per age category at baseline. Lacunar infarcts, cerebral microbleeds and Fazekas scores^23^ of white matter hyperintensities are reported. The Fazekas score is a visual ordinal rating system for white matter hyperintensity severity. It ranges from 0-3, with higher scores indicating more WMH, and is scored separately for periventricular and subcortical regions. The mean ± standard deviation and maximum Fazekas scores for both periventricular and subcortical regions have been included. Summarized are 321 baseline scans; 4 scans were excluded due to poor scan quality or incomplete acquisition.

| Age (y) | Chronic Lacunar Infarcts |  | Cerebral Microbleeds |  | White Matter Hyperintensities (Fazekas Score) |  |  |  |
| --- | --- | --- | --- | --- | --- | --- | --- | --- |
|  | Prevalence | Average per Person | Prevalence | Average per Person | Perivent Prevalence | Perivent; max score | Subcort Prevalence | Subcort; max score |
| **18 - 29** | 1/69<br>(1.4%) | 1.0 | 1/69<br>(1.4%) | 1.0 | 19/67<br>(28.4%) | 0.25 ± 0.45; 1 | 13/67<br>(19.4%) | 0.37 ± 0.45; 2 |
| **30 - 39** | 0/54<br>(0%) | 0.0 | 0/54<br>(0%) | 0.0 | 28/56<br>(50.0%) | 0.47 ± 0.50; 1 | 25/56<br>(44.6%) | 0.55 ± 0.50; 1 |
| **40 - 49** | 0/44<br>(0%) | 0.0 | 2/44<br>(4.5%) | 1.0 | 31/46<br>(67.4%) | 0.64 ± 0.47; 1 | 27/46<br>(57.4%) | 0.74 ± 0.50; 1 |
| **50 - 59** | 2/50<br>(4.0%) | 1.0 | 2/50<br>(4.0%) | 2.0 | 41/48<br>(85.4%) | 0.83 ± 0.39; 2 | 35/48<br>(72.9%) | 0.88 ± 0.51; 2 |
| **60 - 69** | 6/63<br>(9.5%) | 1.3 | 2/63<br>(3.2%) | 1.0 | 51/62<br>(82.3%) | 0.96 ± 0.55; 3 | 49/62<br>(79.0%) | 1.12 ± 0.56; 2 |
| **$\geq 70$** | 9/43<br>(20.9%) | 1.2 | 8/43<br>(18.6%) | 1.5 | 42/42<br>(100.0%) | 1.30 ± 0.62; 3 | 41/42<br>(97.6%) | 1.24 ± 0.54; 3 |

Perivent—periventricular white matter; Subcort—subcortical white matter

Examples of processed quantitative images and maps are shown in Figure 3.

**Figure 3:**
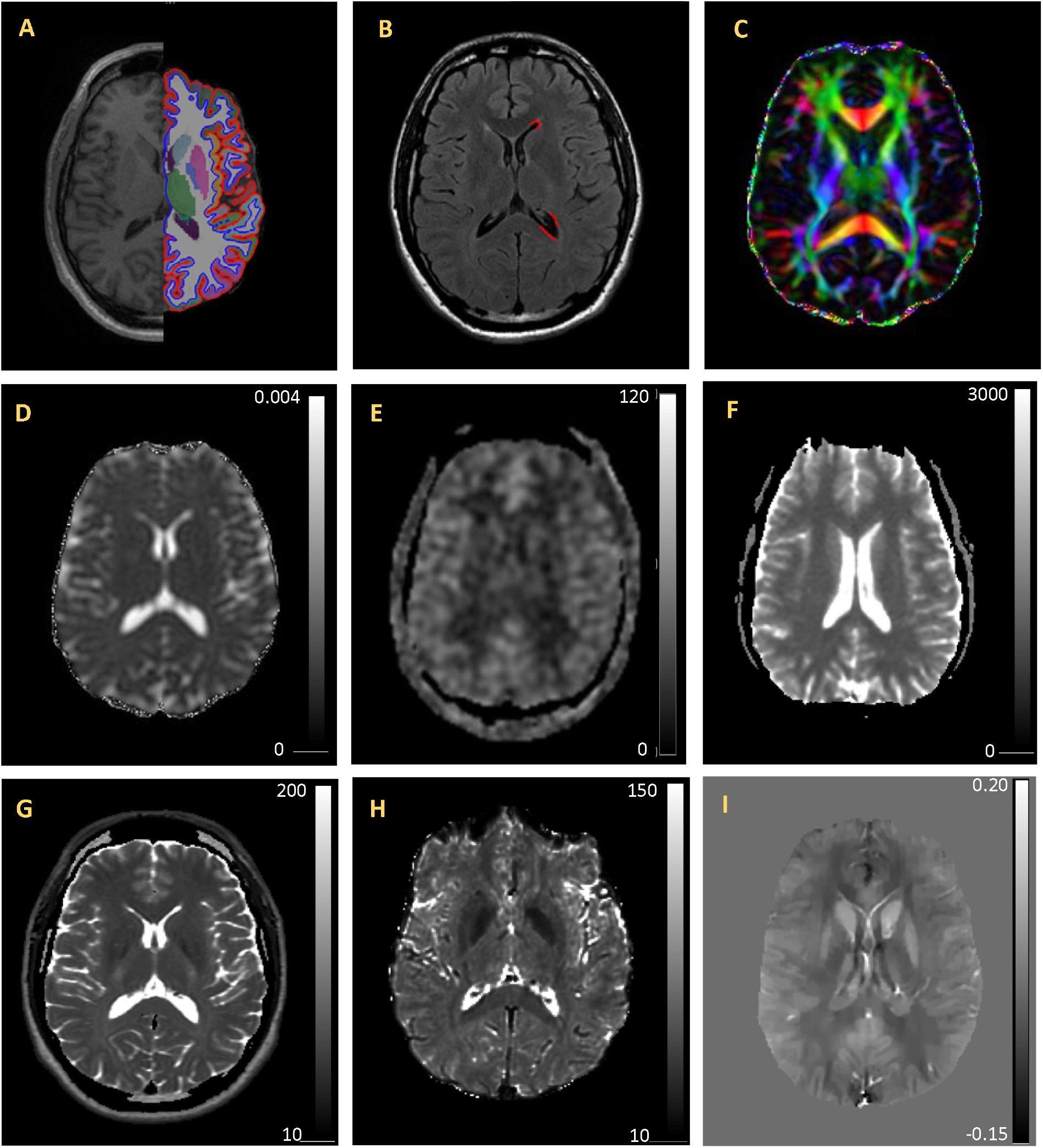
Example quantitative imaging maps and associated processing overlays. A) 3D-T1 image shown with FreeSurfer segmentation and parcellation results overlay on the left hemisphere. B) FLAIR with WMH mask (red) on the left hemisphere C) FA color map indicating the primary diffusion direction of white matter tracts (red indicates left/right; green indicates anterior/posterior; and blue indicates inferior/superior). D) mean diffusivity (MD) map with voxel intensity values are in mm^2^ /s. E) Cerebral blood flow map. Voxel intensity values are in mm/min/100g tissue. F) Quantitative T1 map. Voxel intensity values are in ms. G) T2 map Voxel intensity values are in ms. H) T2* map. Voxel intensity values are in ms. I) Quantitative susceptibility map. Voxel intensity values are in parts per million (ppm). All images are from the same individual and are shown in radiological orientation.

### Protocol Revisions

In the first phase of the study (Table 2A), 3D-T1 images and qT2 were acquired with coronal slice orientation for consistency with other existing research protocols at our centre. Driven equilibrium single observation of T1 (DESPOT1) method18 was implemented. QSM acquisition used unipolar gradients.^19^ After 54 participants were recruited and imaged, the data were reviewed and it was decided to modify the protocol prior to commencing the second phase (Table 2B). Phase 2 protocol adjustments included changing the slice orientation for the 3D-T1 and qT2, implementing a different approach for qT1, and improving the resolution of ASL. We found the DESPOT1 approach provided inconsistent T1 maps with values outside those reported in the literature. A T1 mapping method that is independent of B1 (and thus did not require a B1 map) was developed for brain T1 mapping20. The T2 mapping slice orientation was changed from coronal to axial to harmonize the protocol with newer ongoing studies within our centre. To improve the resolution and reduce through-plane blurring, the acquisition matrix and slice thickness of the ASL sequence were changed. The number of excitations was increased to maintain a similar signal to noise ratio.

### Repeatability and Measurement Validation

To assess short- and long-term repeatability a subset of 1 female and 3 male participants were scanned serially (Figure 1). The mean age (± SD) age of these subjects at time of first scan was 36.6 ± 11.7 years. These four participants have been scanned a total 12 times: 3 scans at each time point (baseline, 18, 23, and 39 months).

Significant changes to our scanner occurred at 1) 9 months into the study (software upgrade version DV22.0 to DV24.0 R01, which included modification of a vendor provided multi-echo spin echo sequence for qT2), 2) 21 months (software and hardware upgrade DV24.0 to DV25 R01), and 3) 33 months (head coil replacement). Other standard service and maintenance changes (repair or replacement of cables, gradient amplifiers, coils, etc.) occurred and are noted in Figure 1. All reported times are from start of the CNS in May 2013. Subsequent analysis of the key quantitative measurements will provide estimates of repeatability, suitable to improve study power calculations.

### Data Sharing

A subset of the imaging and other data has been shared with eight investigators at the University of Calgary. Internal sharing of data was and remains the principle anticipated target group of the CNS Study; however, we also have had opportunity to share data with three investigative teams external to our institution (University of Alberta, Canada; State University of Campinas, Brazil; and University Medical Centre Utrecht, Netherlands). Data equivalent of over 1400 scans have been shared; data from some participants has been shared more than once. The research area of the investigators accessing the CNS data range from depression, inflammatory bowel disease, machine learning, migraine, epilepsy, and stroke. In most cases, only a portion of the acquired data was requested. The most frequently requested imaging data in descending order of demand were 3D-T1, rs-fMRI, FLAIR, pcASL, QSM and qT2.

## DISCUSSION

This report demonstrates a successful recruitment strategy for a prospective, community-recruited quantitative MR study and describes the flexible design and data collection. The data collected will provide estimates of variability and repeatability of each quantitative method and provide data essential for the validation of potential biomarkers of neurodegeneration associated with aging. A distinctive aspect of this study is that it has been designed to incorporate both typical clinical MR sequences and multiple, research-based, quantitative biomarkers including MR relaxometry, diffusion imaging, quantitative susceptibility mapping and cerebral perfusion within a 60-minute scan time. While there are a number of studies that have evaluated one or two of these methods, there are very few that have incorporated this extensive multimodal approach within an imaging session.^10^

### Recruitment and Population Characteristics

The prevalence of neuroimaging findings indicative of covert cerebrovascular small vessels disease in participants in the 70+ age category was up to 20.9%. Similar prevalence of covert lacunar infarcts of presumed vascular origin, white matter hyperintensities and cerebral microbleeds in older community-dwelling populations have been reported previously.^24^ We may need to consider increasing our recruitment numbers in the older age categories to maintain a balanced design of healthy individuals over the adult lifespan. The decision on whether to oversample the older participants is dependent on the definition of what is “normal” aging. Should individuals with possible evidence of cerebral small vessel disease be excluded from the analyses? This question raises some controversy and is as yet unresolved. Given the limited self-reported medical history collected in this study, it will be difficult to determine a definitive diagnosis of cerebral small vessel disease, or other potential confounds like cardiovascular disease.

### MR Protocol

Typically, quantitative MR relaxometry, susceptibility, diffusion and perfusion methods are not all incorporated into either clinical or research studies. This may be due to the additional time required for data acquisition or the expertise and time required for data processing to generate quantitative maps. MR fingerprinting (MRF) has been proposed to overcome a number of these limitation, specifically addressing the longer acquisition times. This emerging approach to quantitative imaging simultaneously generates T1, T2, T2*, and proton density maps from a single acquisition with an approximate scan duration of as little as 16 s.^25 26^ MRF uses voxel-wise pattern matching to a dictionary of MR signal evolution to estimate the MR tissue characteristics. While MRF methods overcome the time limitation for acquisition, these methods are still evolving and have limited availability.

The study design has taken into consideration the evolution of quantitative imaging techniques. Ongoing changes in the MR acquisition protocol can be implemented once an adequate number of participants in each age category have been acquired. The number of participants required can be determined for each quantitative imaging technique. An important outcome of this study will be to establish the measurement variability for each of these quantitative metrics. In practice, a limited number of changes are implemented at any given time. One limitation of this approach is the additional time required for repeatability measures prior to implementation of any intended changes to the MR acquisition protocol.

### Repeatability and Measurement Validation

These repeatability assessments were co-ordinated with selected scheduled changes in the MR operating system. These included major software and hardware upgrades and after the MR system main magnetic field was ramped down for renovations to the MR suite. However, we did not capture repeatability measurements after all unscheduled system changes, including repairs to or replacement of the 12-channel head, neck and spine coil. While this may be a limitation of the repeatability measurements ability to determine the impact of these changes, our MR system is regularly maintained and met the manufacturer specification and operation tolerances after each of these changes. Regular weekly quality assurance and manufacturer service to maintain the MR system within manufacturer specifications is performed, which should mitigate any systems changes.

Another limitation of the repeatability assessment is the limited number and long-term availability of selected participants for evaluation of repeatability and variability. Two participants have recently moved from the local area and new participants have been recruited for future repeatability measures. For purposes of repeatability assessment, we have generally assumed that no significantly physiological changes have occurred in the brains of these participants over either the short or long term. However, if changes were to occur within a given participant, we should be able to identify them as we have nominally three measurements at each time point. These three independent measurements are usually completed within 3 weeks for each individual.

### Data Sharing

Including conventional MR sequences in a purportedly healthy adult population along with research-based sequences, has generated greater interest in data sharing with other local and national studies. It also allows evaluation of these research-based sequences with the context of clinical findings that may help to better understand changes leading to traditional clinical findings. While this study does not match the size or extent of other larger MR initiatives, such as the UK Biobank^27^, we have been able to successfully leverage the data by providing normal control data for other studies.

### Conclusions

Over 55 months from study inception in May 2013, the acquisition protocol has undergone one revision, data for four repeatability time points have been collected, 42-month follow-up data has been collected, and the data has been shared effectively. Our study sample includes ethnically diverse people represented in our local community.^28^ Data collection and analyses remain ongoing. Preparation of detailed reports on repeatability of the quantitative measures, cross-sectional, and longitudinal changes over the adult lifespan are underway.

## Data Availability

MR image data and demographic information are available upon request for academic purposes from qualified researchers. Investigators interested in accessing these data are required to complete a data sharing agreement available on request by emailing. This brief, two-page form, requests a brief description of the study objectives and intended purpose, a list of MR sequences of interest, required medical information, and demographic information. The sharing agreement also requires that the study requesting data has approval from the appropriate REB.

## ACKNOWLEDGEMENTS

This ongoing study would not be possible without assistance from a number of individual trainees, technicians, and physicians including Brie Crawston-Grant, BSc; Qian (Lucy) Lu, MSc; Paul Romo, MRT; Courtney Seefeldt, Robert J Sevick, MD; and Tamara Trainor, MRT.

## AUTHORS’ CONTRIBUTIONS

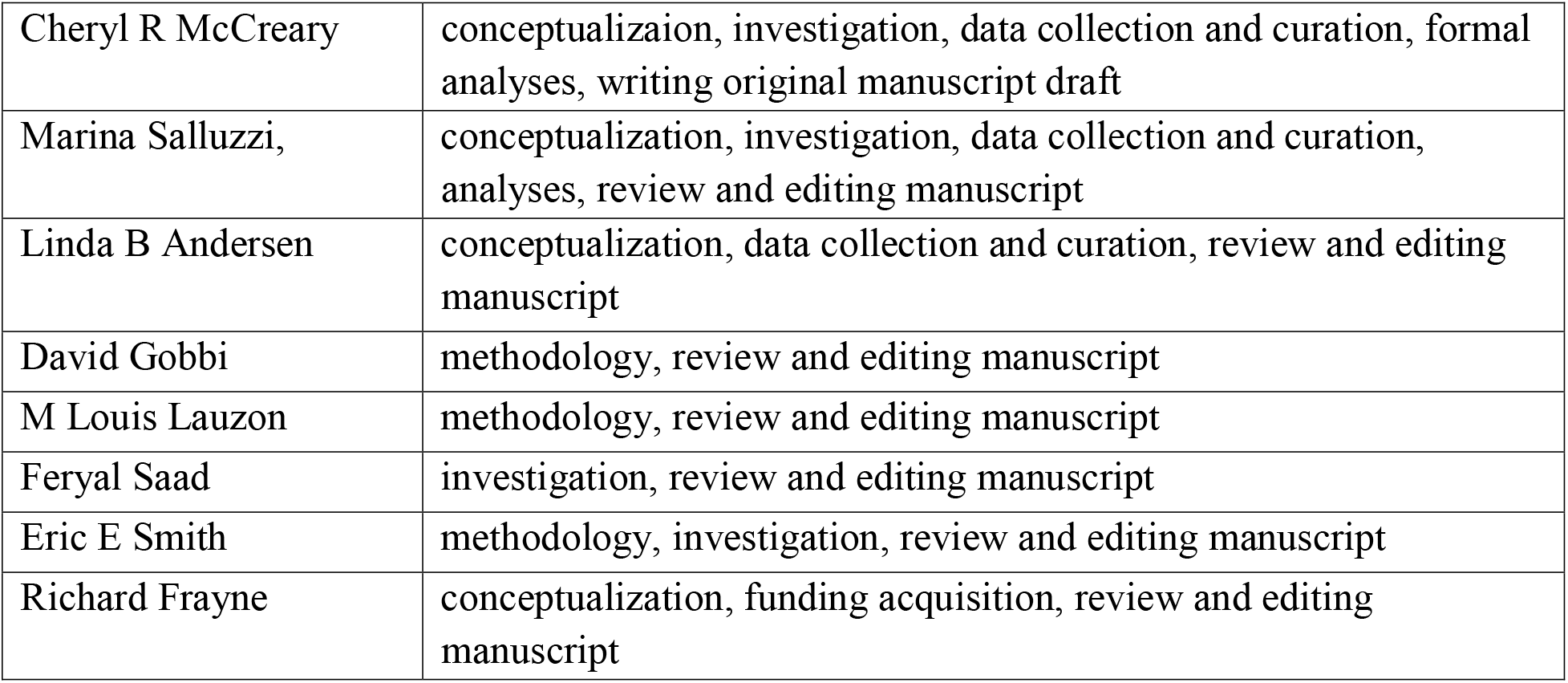

